# Genome-wide Association Study of Non-syndromic Orofacial Clefts in a Multiethnic Sample of Families and Controls Identifies Novel Regions

**DOI:** 10.1101/2020.10.27.20220574

**Authors:** Nandita Mukhopadhyay, Eleanor Feingold, Lina Moreno-Uribe, George Wehby, Luz Consuelo Valencia-Ramirez, Claudia P. Restrepo Muñeton, Carmencita Padilla, Frederic Deleyiannis, Kaare Christensen, Fernando A. Poletta, Ieda M Orioli, Jacqueline T. Hecht, Carmen J. Buxó, Azeez Butali, Wasiu L. Adeyemo, Alexandre R. Vieira, John R. Shaffer, Jeffrey C. Murray, Seth M. Weinberg, Elizabeth J. Leslie, Mary L. Marazita

**Affiliations:** Center for Craniofacial and Dental Genetics, Department of Oral Biology, School of Dental Medicine, University of Pittsburgh, Pittsburgh, PA, 15219 USA; Department of Biostatistics, Graduate School of Public Health, University of Pittsburgh, Pittsburgh, PA, USA; Department of Human Genetics, Graduate School of Public Health, University of Pittsburgh, Pittsburgh, PA, USA; Department of Orthodontics, & The Iowa Institute for Oral Health Research, College of Dentistry, University of Iowa, Iowa City, IA, USA; Department of Health Management and Policy, College of Public Health, University of Iowa, Iowa City, IA, USA; Fundación Clínica Noel; Calle 14 # 43B – 146, Medellín, Antioquia, Colombia; Department of Pediatrics, College of Medicine, Institute of Human Genetics, National Institutes of Health, University of the Philippines, Manila, the Philippines; UCHealth Medical Group, Colorado Springs, CO. USA; Department of Epidemiology, Institute of Public Health, University of Southern Denmark, Odense, Denmark; CEMIC: Center for Medical Education and Clinical Research, Buenos Aires, Argentina; Department of Genetics, Institute of Biology, Federal University of Rio de Janeiro, Rio de Janeiro, Brazil; Instituto Nacional de Genética Médica Populacional INAGEMP, Porto Alegre, Brazil; Department of Pediatrics, University of Texas Health Science Center at Houston, Houston, TX, USA; Dental and Craniofacial Genomics Core, School of Dental Medicine, University of Puerto Rico, San Juan, Puerto Rico; Department of Oral Pathology, Radiology and Medicine, Iowa Institute for Oral Health Research, College of Dentistry, University of Iowa, Iowa City, IA, USA; Department of Oral and Maxillofacial Surgery, College of Medicine, University of Lagos, Lagos, Nigeria; Department of Pediatrics, Carver College of Medicine, University of Iowa, Iowa City, IA, USA; Department of Human Genetics, Emory University, Atlanta, GA, USA; Clinical and Translational Science, School of Medicine, University of Pittsburgh, Pittsburgh, PA, USA

## Abstract

Orofacial clefts (OFCs) are among the most prevalent craniofacial birth defects worldwide and create a significant public health burden. The majority of OFCs are non-syndromic and vary in prevalence by ethnicity. Africans have the lowest prevalence of OFCs (∼ 1/2,500), Asians have the highest prevalence (∼1/500), European and Latin Americans lie somewhere in the middle (∼1/800 and 1/900 respectively). Thus, ethnicity appears to be a major determinant of the risk of developing OFC. The Pittsburgh Orofacial Clefts Multiethnic study was designed to explore this ethnic variance, comprising a large number of families and individuals (∼12,000 individuals) from multiple populations worldwide: US and Europe, Asians, mixed Native American/Caucasians, and Africans. In this current study, we analyzed 2,915 OFC cases, 6,044 unaffected individuals related to the OFC cases, and 2,685 controls with no personal or family history of OFC. Participants were grouped by their ancestry into African, Asian, European, and Central and South American subsets, and genome-wide association run on the combined sample as well as the four ancestry-based groups. We observed 22 associations to cleft lip with or without cleft palate at 18 distinct loci with p-values < 1e-06, including 10 with genome-wide significance (< 5e-08), in the combined sample and within ancestry groups. Three loci - 2p12 (rs62164740, p=6.27e-07), 10q22.2 (rs150952246, p=3.14e-07), and 10q24.32 (rs118107597, p=8.21e-07) are novel. Nine were in or near known OFC loci - *PAX7, IRF6, FAM49A, DCAF4L2*, 8q24.21, *NTN1, WNT3-WNT9B, TANC2*, and *RHPN2*. The majority of the associations were observed only in the combined sample, European, and Central and South American groups. We investigated whether the observed differences in association strength were a) purely due to sample sizes, b) due to systematic allele frequency difference at the population level, or (c) due to the fact certain OFC-causing variants confer different amounts of risk depending on ancestral origin, by comparing effect sizes to observed allele frequencies of the effect allele in our ancestry-based groups. While some of the associations differ due to systematic differences in allele frequencies between groups, others show variation in effect size despite similar frequencies across ancestry groups.

## 1 Introduction

Orofacial clefts are among the most common birth defects in all populations worldwide and pose a significant health burden. Surgical treatment along with ongoing orthodontia, speech and other therapies, are very successful in ameliorating the physical health effects of OFC, but there is still a significant social, emotional and financial burden for individuals with OFC, their families, and society (Wehby and Cassell, 2010; Nidey et al., 2016). Furthermore, there are disparities in access to such therapies for OFCs (Nidey and Wehby, 2019), similar to other malformations with complex medical and surgical needs. Some studies have suggested a reduced quality of life for individuals with OFCs (Naros et al., 2018), while other studies have identified higher risk to certain types of cancers (Bille et al., 2005; Taioli et al., 2010; Bui et al., 2018). Thus, it is critical to identify etiologic factors leading to OFCs to improve diagnostics, treatments, and outcomes.

OFCs manifest themselves in many forms - cleft lip alone (CL), cleft palate alone (CP), combination of the two (CLP), and vary in their severity. A large proportion of the causal genes involved with syndromic OFCs (i.e. OFCs that are part of other syndromes) are known (OMIM, https://www.omim.org/search/advanced/geneMap). However, the majority of OFC cases - including about 70% of CL with or without CP (CL/P) and 50% of CP alone - are considered non-syndromic, i.e. occurring as the sole defect without any other apparent cognitive or structural abnormalities (Dixon et al., 2011). The genetic causes of non-syndromic OFCs are still largely undiscovered.

There are differences in birth prevalence around the world with respect to OFC, that may be the result of etiological differences among these different populations, both genetic and non-genetic. Populations of Native American and Asian ancestry have the highest prevalence of approximately 2 per 1,000 live births, European ancestry populations have an intermediate prevalence of ∼1 per 1,000, and African-ancestry populations have the lowest prevalence, ∼ 1 per 2,500 (Dixon et al., 2011). Most of the reported GWAS loci are observed within specific ancestry groups, and there are only a few studies that conducted a systematic comparison of genetic differences between populations of different ancestry, largely due to the lack of sufficiently large samples.

In the current study, we expanded this analysis to other loci and additional populations from the Pittsburgh Orofacial Clefts (POFC) study to describe differences in the genetic etiology of OFC across populations of varying ancestral origin. The POFC Multiethnic study is a geographically diverse, family-based study comprising a large number of families and individuals (∼12,000 participants) from multiple populations worldwide including those of European ancestry from the US and Europe, Asians from China and the Philippines, mixed Native American, European and African ancestry from Central and South America, and Africans from Nigeria and Ethiopia. The POFC Multiethnic study sample is therefore a rich resource for examining the genetic heterogeneity across populations. In two previous GWAS studies (Leslie et al., 2016; Leslie et al., 2017), a subset of the total POFC participants were analyzed, including 1,319 independent parent-offspring trios, 823 unrelated cases and 1700 controls. These previous GWASs have focused on unrelated cases and controls, or parent-offspring trios, not fully taking advantage of the fact that OFCs often segregate within families. Here, we include all available participants from simplex and multiplex pedigrees, along with singleton cases and controls for a total sample size of 2,915 affected and 8,729 unaffected individuals. As a result of the increased sample size, our study should provide more power to detect novel OFC loci, while providing stronger evidence for previously reported associations that are common to all OFCs.

## 2 Materials and methods

### 2.1 Study Sample

The current study sample consists of 2,915 participants with OFCs, 6,044 unaffected individuals related to the OFC cases, and 2,685 controls with no personal or family history of OFC. The participants were recruited from multiple sites from Asia (China, India, Philippines and Turkey), Europe (Spain, Hungary, Denmark), Africa (Nigeria and Ethiopia) and the Americas (Argentina, Colombia, Guatemala, mainland USA and Puerto Rico). Genotyping was performed at the Center for Inherited Disease Research (CIDR) at Johns Hopkins University, on the Illumina platform on approximately 580 K variants. QC was carried out by the CIDR genotyping coordination center at the University of Washington. Genotypes were then imputed using the 1000 genome project phase 3 reference panel, at approximately 35,000,000 variants of the GrCH37 genome assembly. More details on the POFC Multiethnic study can be found in the Database of Genotypes and Phenotypes (dbGaP; study accession number phs000774.v2.p1) and at the FaceBase resource for craniofacial research under dataset OFC4: Genetics of Orofacial Clefts and Related Phenotypes (URL https://www.facebase.org/id/1-50DT) (Mary L. Marazita, 2019).

Ancestry of a subset of unrelated participants from the POFC Multiethnic study was determined based on principal components using genotyped variants, which were then projected onto the larger sample. The first three principal components of ancestry correspond well with the broad geographical origin of the POFC participants, except for those from United States, and were used to classify participants as being of African, Asian, European, or Central and South American origin; this same classification was used for group analyses. Participants from the United States were included in African, European and Central and South American ancestry groups. Participants from Turkey are part of the European ancestry group. A more detailed description of ancestry determination can be found in the previously published GWASs of POFC Multiethnic study participants (Leslie et al., 2016; Mary L. Marazita, 2019). The first three PCs provide a workable, although not perfect, separation of participants into the four broad ancestry groups within the POFC Multiethnic study sample. Note that the PCs were not used to account for population substructure in the actual association analysis, therefore we did not need to utilize the higher order PCs.

Genome-wide association was run on the combined study sample (**ALL**), as well as the four ancestry-based groups, Africans (**AFR**), Asians (**ASIA**), Europeans (**EUR**), and Central and South Americans (**CSA**). The sample sizes for each ancestry-based group are provided in Table 1. Table 1 contains the median and maximum pedigree sizes, as well as the median and maximum number of affected individuals per pedigree. The AFR group contains mainly singleton CL/P cases and controls, whereas ASIA and EUR have a larger proportion of extended families; the CSA group’s pedigree sizes lie somewhere in between.

**Table 1.** Study sample characteristics

|  | **AFR** | **ASIA** | **EUR** | **CSA** | **ALL** |
| --- | --- | --- | --- | --- | --- |
| Total pedigrees | 227 | 484 | 1,271 | 1,433 | 3,422 |
| CL/P pedigrees | 152 | 320 | 507 | 944 | 1,925 |
| Median CL/P pedigree size | 1 | 1 | 1 | 1 | 3 |
| Maximum CL/P pedigree size | 9 | 34 | 35 | 27 | 35 |
| Unaffected pedigrees | 75 | 164 | 764 | 489 | 1,498 |
| Median unaffected pedigree size | 1 | 1 | 1 | 3 | 1 |
| Maximum unaffected pedigree size | 5 | 4 | 15 | 21 | 21 |
| Total individuals | 428 | 1,691 | 3,273 | 4,038 | 9,447 |
| Total unaffected individuals | 274 | 1,246 | 2,704 | 2,988 | 7,226 |
| Total affected individuals | 154 | 445 | 569 | 1,050 | 2,221 |
| Median number of affected members per CL/P pedigree | 1 | 1 | 1 | 1 | 1 |
| Maximum number of affecteds per CL/P pedigree | 2 | 5 | 5 | 4 | 5 |

### 2.2 Phenotype definition

In our study, we ran GWAS of CL/P on participants from pedigrees in which the OFC-affected members have a cleft lip only (CL), or a cleft lip and a cleft palate (CLP). Pedigrees with members who are affected with a cleft palate only, or a reported history of cleft palate only, were excluded from our analysis. Within these pedigrees, any member with an OFC was considered to be affected for CL/P; unaffected pedigree members from CL/P pedigrees as well as control pedigrees that do not have any history of OFCs were considered to be unaffected for CL/P. The final combined sample (ALL) included 2,221 affected and 7,226 unaffected participants.

### 2.3 Statistical method

The program EMMAX (Kang et al., 2010) was used to run the seven GWAS. EMMAX uses a variance component mixed model approach to detect association at each variant, while accounting for population structure and familial relatedness. Therefore, ancestry PCs are not included in the association analysis. Instead, a genetic relationship matrix (GRM) estimated from the observed genotype data is used within the linear mixed model. The genetic relationship matrix is also used to estimate the polygenic variance component. Strength of association is then measured using a score test of comparison between the maximum likelihood conditional on observed genotypes at each variant versus the unconditional maximum likelihood model. Due to the nature of the score test, the reported effect size is not interpretable for a binary phenotype, therefore we do not utilize the effect sizes reported by EMMAX. Instead, another mixed-model association program, GENESIS (Gogarten et al., 2019) was used to calculate approximate effect sizes (betas and standard errors). The same effect allele is used to report effect size and direction consistently across groups, namely the minor allele at each variant as identified in the combined POFC sample.

### 2.4 Filtering of variants and identification of associated loci

We included variants that had minor allele frequencies of 2% or more within their respective ancestry groups. In this study, loci containing variants with association p-value less than 1.0e-06 were reported as positive associations. The observed minor allele frequencies of reported loci were compared to the respective population frequencies from the gnomAD database (Karczewski et al., 2019) to ensure that minor allele frequencies were not impacted by the imputation. Note that major and minor alleles may be switched between groups, but this impacts only the direction of effect, not the strength of association.

### 2.5 Identification of novel OFC loci

We compared significant and suggestive associations from our GWASs to genes and regions that have been identified through genome-wide linkage or association, as well as by candidate gene studies by multiple previous genetic studies of OFCs. Our reference loci consist of the 29 genomic regions listed in the review article by Beaty et al. (Beaty et al., 2016), combined with loci reported by six recently published GWASs. The six newer GWASs include (1) combined meta-analysis of parent-offspring trio and case-control cohorts from the current POFC multiethnic study sample (Leslie et al., 2016), (2) meta-analysis of the cohorts used in (1) with another OFC sample consisting of European and Asian participants (Leslie et al., 2017), (3) GWAS of cleft lip with cleft palate in Han Chinese samples (Yu et al., 2017), (4) GWAS of cleft lip only and cleft palate only in Han Chinese (Huang et al., 2019), (5) GWAS of cleft lip with or without cleft palate in Dutch and Belgian participants (van Rooij et al., 2019), and (6) GWAS of sub-Saharan African participants from Nigeria, Ghana, Ethiopia and the Republic of Congo (Butali et al., 2019).

For each comparison, we derived search intervals for detecting overlaps between published/known OFC loci and the associations reported in this current study. For each known OFC gene, GrCH37 base pair positions of start and end transcription sites from the UCSC genome browser were used to measure distance between our locus and the gene. For the 8q24.21 locus, which is a gene desert, we checked whether any of our associated SNPs were located within the 8q24.21 chromosome band. A positive replication is reported where the minimum p-value observed within 500 KB on either side of the gene exceeds 1e-05. For comparing published GWAS loci, separate 1 MB search intervals were created around the reported lead variants, and a positive replication noted if any of our selected associations were within 500 KB of these intervals.

### 2.6 Comparison between ancestry groups

For comparing GWASs of ancestry groups, we first compared association results, namely, effect size and direction, and association p-value of each lead variant within each ancestry group, as well as variants nearby. However, comparing association outcomes at a single variant is not always feasible, nor advisable, as association outcomes are impacted by differences in variant allele frequencies and LD, which differ by ancestry. We therefore looked at intervals of 500 KB to either side of the lead SNPs, and selected the 30 top associations observed within these intervals within each GWAS. Approximate effect sizes for selected variants were computed using the same effect allele at each variant, which is the minor allele for that variant in ALL. GENESIS (Gogarten et al., 2019) reports the beta coefficient for each SNP in terms of allele dosage. In our analysis, a positive effect size indicates that the effect allele increases risk of CL/P and vice-versa.

We hypothesized that the main reason for difference in association outcomes between ancestry groups is due to the fact that variants are common in one population, but rare in others. If an OFC causing variant is observed with similar frequency in multiple groups, we expect to see elevated association signals in all these groups as well. Although the strength of association indicated by a p-value is dependent on sample size and may be reduced in the smaller groups, the magnitude and direction of effect sizes should be similar. If this is not the case, it implies that variant’s contribution to the development of OFCs varies depending on the participants’ ancestral origin. Thus, we compared the effect size and effect allele frequencies of the top 30 associations observed close to each lead SNP. In order to assess whether allele frequencies are different between groups, we calculated the fixation index F _ST_ for the top 30 associations, using the ancestry-based groups in our ALL sample, and used the 95% percentile of the distribution of F _ST_ as a threshold value to decide whether the effect allele frequencies were similar or different. To compare effect size estimates, we examined whether the point estimate was in the same direction and if the confidence intervals overlapped. We also ran a meta-analysis of the selected variants as a statistical means for measuring whether the effect allele affects CL/P risk similarly across ancestry groups. The meta-analysis procedure reports the p-value for Cochrane’s Q statistic at each analyzed variant, testing the hypothesis that effect sizes differ across the groups. PLINK (Chang et al., 2015) was used to run meta-analysis and for calculation of Wright’s F _ST_ fixation index.

We used these comparisons to categorize each locus into one of three categories:

Category 1: The effect alleles have similar frequencies within the groups, and association p-values are estimates are also similar.

Category 2: Effect allele frequency differs across groups, effect size estimates may or may not differ.

Category 3: Allele frequencies are similar between groups, but the effect size estimates differ.

## 3 Results

GWASs of the combined sample and the four ancestry-based groups yielded 22 associations with p-values less than 1e-06, including 10 genome-wide significant associations (i.e. below the Bonferroni threshold of 5e-08) at 18 distinct loci. In some cases, these loci are observed in more than one ancestry-based group (Figure 1, Table 2). Of the 18 loci, 15 are in or near reported OFC genes and reported associations from the published OFC GWASs listed in **Methods**, and the other three are novel. The novel associations include (i) chromosome 2p12 (rs62164740, p-value 6.27E-07); (ii) 10q22.2 (rs150952246, p-value 3.14E-07); and (iii) 10q24.32 (rs118107597, p-value 8.21E-07). All three showed the strongest association in the combined ALL sample. Four of the 22 associations were observed within a specific ancestry group: these included (i) chromosome 9q33.1 in EUR, rs9408874, p-value 2.59E-07; (ii) 12q13.2 in CSA, rs34260065, p-value 3.87E-07; (iii) *TANC2* in EUR, rs1588366, p-value 1.09E-08; and (iv) 17q25.3 in AFR, rs1975866, p-value 3.13E-08. The ASIA subgroup did not yield any association with a significance p-value less than our threshold of 1e-06. The strongest associations were seen at the 8q24 locus with p-value 2.80E-29 in ALL, as well as exceeding genome-wide significance in EUR and CSA. Table 2 lists the 18 associations, with the three novel loci are listed in bold. Figure 1 shows the Manhattan plots of the five GWASs.

**Table 2.** Associations with p values < 1e-06

| Locus | Chromosome band | Lead SNP (effect allele) | Lead SNP position | P value<sup>A</sup> | Effect size, Std. error<sup>B</sup> | Groups with p < 1e-06<sup>A</sup> | OFC Gene/prior GWAS |
| --- | --- | --- | --- | --- | --- | --- | --- |
| 1 | 1p36.13 | rs9439714 (C) | 18,976,489 | 3.27E-08 | 0.15 ± 0.04 | ALL | *PAX7* |
| 2 | 1q32.2 | rs17015217 (A) | 209,966,629 | 1.46E-09 | -0.19 ± 0.05 | ALL, CSA | *IRF6* |
| 3 | 2p24.3-24.2 | rs6745357 (G) | 16,713,395 | 3.62E-10 | 0.25 ± 0.03 | ALL | *FAM49A* |
| **4** | **2p12** | **rs62164740 (A)** | **78,177,488** | **6.27E-07** | **0.25 ± 0.06** | **ALL** |  |
| 5 | 5p13.2-13.1 | rs181764204 (T) | 38,205,024 | 2.63E-07 | 0.54 ± 0.10 | ALL | (Huang et al., 2019) |
| 6 | 5q35.2-35.1 | rs17075892 (T) | 172,827,353 | 6.95E-08 | -0.25 ± 0.04 | ALL | (Butali et al., 2019; Huang et al., 2019) |
| 7 | 8q21.3 | rs12543318 (C) | 88,868,340 | 7.45E-11 | 0.24 ± 0.04 | ALL, EUR | *DCAF4L2* |
| 8 | 8q22.2 | rs185266751 (G) | 99,712,644 | 2.49E-07 | 0.50 ± 0.09 | ALL | (Huang et al., 2019) |
| 9 | 8q24.21 | rs72728755 (A) | 129,990,382 | 2.80E-29 | 0.48 ± 0.05 | ALL, CSA, EUR | 8q24 |
| 10 | 9q33.1 | rs9408874 (T) | 118,279,489 | 2.59E-07 | 0.26 ± 0.05 | EUR | (Huang et al., 2019) |
| **11** | **10q22.2** | **rs150952246 (C)** | **77,151,597** | **3.14E-07** | **0.37 ± 0.08** | **ALL** |  |
| **12** | **10q24.32** | **rs118107597 (A)** | **104,283,510** | **8.21E-07** | **0.58 ± 0.12** | **ALL** |  |
| 13 | 12q13.2 | rs34260065 (C) | 55,540,318 | 3.87E-07 | 0.12 ± 0.04 | CSA | (Huang et al., 2019) |
| 14 | 17p13.1 | rs16957821 (G) | 8,948,104 | 9.12E-10 | 0.25 ± 0.04 | ALL | *NTN1* |
| 15 | 17q21.31-21.32 | rs7216951 (T) | 44,972,810 | 1.42E-07 | -0.20 ± 0.04 | ALL | *WNT3, WNT9B* |
| 16 | 17q23.2-23.3 | rs1588366 (G) | 61,076,428 | 1.09E-08 | -0.24 ± 0.05 | EUR | *TANC2* |
| 17 | 17q25.3 | rs1975866 (C) | 77,267,377 | 3.13E-08 | 0.39 ± 0.09 | AFR | (Huang et al., 2019) |
| 18 | 19q13.11 | rs2003950 (A) | 33,546,283 | 1.21E-07 | 0.21 ± 0.04 | ALL | *RHPN2* |

Note: Novel loci highlighted in bold; <sup>A</sup>Association p-values from the EMMAX program; <sup>B</sup>Effect size (Beta coefficient) and standard error of effect size from the GENESIS program.

**Figure 1.**
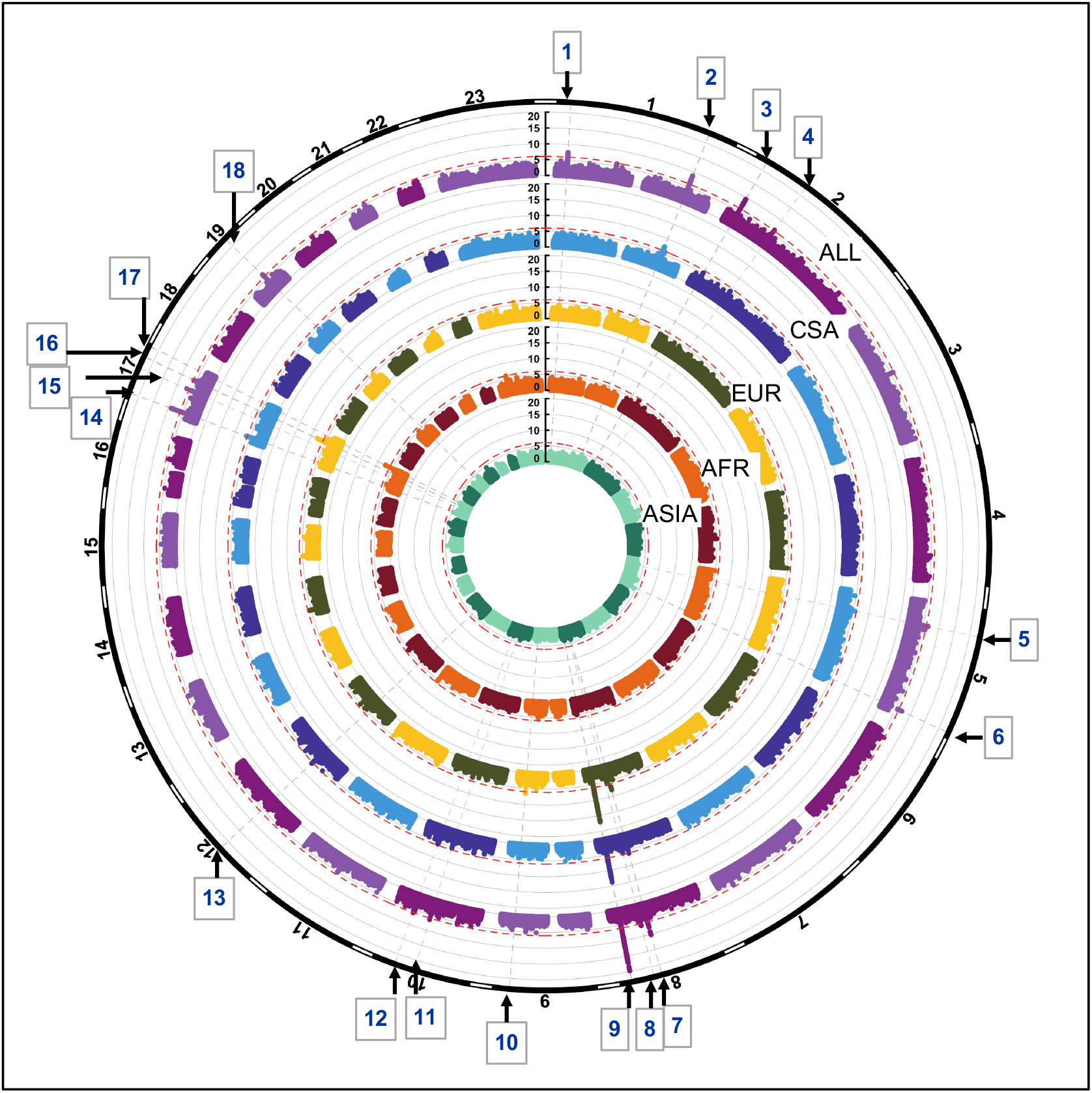
Circular Manhattan plots of ALL, CSA, EUR and ASIA. Red lines are drawn at the 1e-06 significance level. Loci significant at the 1e-06 significance level are indicated by the gray dotted lines and numbered 1-18. Chromosomes are depicted by the black bands in the outermost circle.

### 3.1 Novel associations and replication

There were three novel associations that did not correspond to known OFC GWAS loci, including the previously published GWASs of POFC Multiethnic Study participants (Leslie et al., 2016; Leslie et al., 2017), GWAS of Europeans (van Rooij et al., 2019), and GWAS of Africans (Butali et al., 2019). Along with the three novel loci, we also investigated four additional associations that were reported in Han Chinese GWASs with significance below the genomewide suggestive threshold of 1e-05 for possible roles in the development of CL/P. Interestingly, the four loci reported in Chinese samples do not show association with the ASIA group in our study, rather they are observed with ancestry groups other than Asians in our current study. These 7 loci are listed in Table 3 along with the gene nearest the lead SNP. The nearest gene was identified based on gene locations, specifically, transcription start and end positions obtained from the UCSC genome browser. In Table 3, we also note whether there are regulatory elements such as super-enhancers involved in craniofacial development, or if these regions appear to be expressed in fetal craniofacial tissue, as listed in Epigenomic Atlas of Early Human Craniofacial Development (Wilderman et al., 2018; Justin Cotney, 2020).

**Table 3.** Novel loci, closest gene and contribution to craniofacial development

| **Locus** | **Chrom. band** | **Groups with $p < 1e-06$** | **Nearest Gene** | **Regulatory elements for craniofacial development and other roles in OFCs** |
| --- | --- | --- | --- | --- |
| **4** | 2p12 | ALL, CSA | *SNAR-H* | Evidence of methylation during craniofacial development |
| 8 | 8q22.2 | ALL, CSA | *STK3* | Transcription start site, upstream promoter and enhancer region |
| 10 | 9q33.1 | EUR | *LINC00474* | Craniofacial super-enhancer region |
| **11** | 10q22.2 | ALL, CSA | *ZNF503* | Transcription start site and promoter region |
| **12** | 10q24.32 | ALL, CSA | *SUFU* | Promoter, transcription enhancer; *SUFU* is involved in the Hedgehog signaling pathway |
| 13 | 12q13.2 | CSA | *OR9K2* | Craniofacial super-enhancer region; *OSR2* gene 240Kb upstream codes for transcription factor shown to control palatal development (Lan et al., 2004; Fu et al., 2017) |
| 17 | 17q25.3 | AFR | *RBFOX3* | Craniofacial super-enhancer region; *TIMP2* gene 335Kb connected to increased risk of CL/P (Letra et al., 2014) |

Note: Peak numbers in bold indicate novel loci, other loci showed modest association ( $p < 1e-04$ ) in GWAS of Chinese (Huang et al., 2019)

**Table 4.** Comparison of effect allele frequency and effect size of lead SNPs within each locus

| Peak | Locus | Gene | Category | P-value of Q statistic | Effect allele frequency | Main SNP effect (Beta) of top SNPs |
| --- | --- | --- | --- | --- | --- | --- |
| 1 | 1p36.13 | *PAX7* | 2 | Not significant | Low in ASIA | Smaller in ASIA at several SNPs |
| 2 | 1q32.2 | *IRF6* | 2 | Not significant | Low in EUR, AFR | Larger in EUR than CSA, ASIA |
| 3 | 2p24.3-24.2 | *FAM49A* | 2 | Not significant | Effect allele common in all groups | Similar effect size estimates at most SNPs |
| 4 | 2p12 |  | 1 | Not significant | Low in ASIA | Larger in ASIA but not statistically different |
| 5 | 5p13.2-13.1 |  | 1+3 | p(Q) < 0.05 at some SNPs, | Low in EUR, similar in others | Larger in EUR at SNPs with significant p(Q) |
| 6 | 5q35.2-35.1 |  | 1 | Not significant | Similar across groups | More negative in AFR, but not statistically different |
| 7 | 8q21.3 | *DCAF4L2* | 2+3 | Highly significant at a few SNPs, p(Q) < 0.001 | Similar across groups | Smaller in ASIA at SNPs with p(Q) < 0.001 |
| 8 | 8q22.2 |  | 2 | Not significant | Common (15%-50%) in CSA, a few variants rare in others (< 2%). | Similar across groups |
| 9 | 8q24.21 |  | 1 | Not significant | Low in ASIA (2%-5%) | Similar across groups |
| 10 | 9q33.1 |  | 3 | Significant | Similar across groups | Significantly larger in EUR |
| 11 | 10q22.2 | | 2 | Not significant except a few SNPs with p(Q) ≈ 0.05 | Rare in AFR (< 2%) | Larger in AFR at SNPs at SNPs with p(Q) ≈ 0.05 |
| 12 | 10q24.32 |  | 1 | Not significant | Low in all groups (< 2% - 15%) | Similar across groups |
| 13 | 12q13.2 |  | 3 | p(Q) < 0.05 | Common in all groups (15% - 45%) | Significantly larger in CSA, larger but not significantly so in AFR |
| 14 | 17p13.1 | *NTN1* | 1 | Not significant except one, p(Q) < 0.01 | Common in all groups | Significantly larger in AFR only for SNP with significant p(Q) |
| 15 | 17q21.31-21.32 | *WNT3*,<br>*WNT9B* | 1 | Not significant | Common in all groups (15% - 45%) | Similar across groups |
| 16 | 17q23.2-23.3 | *TANC2* | 3 | Significant, p(Q) < 0.01 | Low in ASIA (5%-15%) | Significantly larger in EUR |
| 17 | 17q25.3 |  | 3 | Significant, p(Q) < 0.001 | Common in all groups (> 15%) | Significantly larger in AFR |
| 18 | 19q13.11 | *RHPN2* | 2 | Not significant | Common in all groups | Similar effect sizes |

### 3.2 Comparison of associations by ancestry group

Consistent with previous studies, we observed that many loci show significant association in one ancestry group, but are well below the suggestive threshold in the others. We wanted to explore the possible explanations for these findings and categorized each locus into one of three categories, which we describe below. We were able to easily categorize some of the associated loci into one of these three groups, while others were not as definitive. Table 1 lists the category that best fits each associated locus and summarizes the characteristics of top associations at each locus in brief, with respect to their allele frequencies, main SNP effects, and significance of the Q-statistics. The effect sizes are noted as being different if their confidence intervals do not overlap, and allele frequencies are considered to vary across groups if the F_ST_ value falls above 0.065, which is the 95^th^ percentile of the genome-wide distribution of F_ST_ values in ALL. From these results, one can conclude that consistently significant Q statistics within a locus is indicative of a category 3 locus (when allele frequencies are similar), wherein the same variant contributes differently to CL/P risk in populations with different ancestry. The reverse is almost always true where the variant has similar effects irrespective of ancestry, i.e. those located within category 1 loci. Category 2 loci appear to lie somewhere in between.

#### Category 1

The significance of association appears to be a function of the sample size. In this category, the effect sizes are similar in all four ancestry groups; therefore, larger ancestry groups (e.g., CSA) and/or larger effect allele frequencies will have greater power to detect an association than the smaller ancestry groups (e.g., Asian and African), and/or if the effect allele is rare. We found this to be the case for six loci, which included 2p12, 5q35.1-q35.2, 8q24.21, 10q24.32 (novel locus), 17p13.1 (*NTN1*) and 17q21.31-q21.32 (*WNT3* and *WNT9B*). The Q statistic p-value for meta-analysis is not significant at the nominal threshold of 5% at the top associated positions, and F_ST_ values are generally small (0.05 or less). The 8q24.21 locus is a special case where effect allele frequencies of top associated SNPs are much lower in the ASIA group than the others.

#### Category 2

Association p-values differ between groups due to the fact that the allele frequencies are different. Effect size and direction appear to be similar across the subgroups. Six associated loci, 1p36.13 (*PAX7*), 1q32.2 (*IRF6*), 2p24.3-p24.2 (*FAM49A*), 8q22.2, 10q22.2, and 19q13.11 (*RHPN2*) are consistent with this category. The Q statistic is not significant, however, F_ST_ values are on the average high (> 0.1).

#### Category 3

Association p-values differ between groups. Effect sizes and direction vary substantially indicating that the same allele increases risk of OFC in certain ancestry groups, but has the opposite effect or has no effect on risk of OFC in others. However, effect allele frequency match across groups. Four associations, 9q33.1, 12q13.2, 17q23.2-q23.3 (*TANC2*), and 17q25.3 are consistent with this category.

#### Uncertain category

The 5p13.2-p13.1 locus appears to contain associated SNPs from categories 1 and 3. The effect sizes are not the same for all groups at a few SNPs, however, effect allele frequencies appear to be mostly similar, with F_ST_ values around 0.05. The 8q21.3 (*DCAF4L2*) locus also contains a few loci that belong in category 3, where effect allele frequencies are the same across all groups (F_ST_ < 0.05), while effects differ significantly (p-value of Q statistic < 1e-03). Other variants conform to category 2, where the effect sizes are statistically similar, but allele frequencies differ.

An example from each of the three categories are shown in figures 2-4 below, the 17p13.1 (*NTN1*) locus from category 1, the 1q32.2 (*IRF6*) locus from category 2, and the novel association in AFR at 17q25.3 from category 3. Each figure shows a regional plot of association p-values (1^st^ row), effect size and confidence intervals (forest plot in 2^nd^ row left), and allele frequencies of the effect allele in each group (heatmap in 2^nd^ row right). The regional plot spans the top 30 associations observed at that locus, with the top 10 SNPs identified. The forest plots show effect sizes and confidence intervals on the effect sizes (x axis) of the top 10 SNPs (listed on the y-axis) for each group. The Q-statistic p-value (precision above 0.001) is noted below each variant in the forest plots. The heatmap shows effect allele frequencies of the top 10 variants for each group (y-axis); the lead variant is highlighted in red within each heatmap. Variants are ordered by base-pair position from top to bottom in each forest plot of effect size, and allele frequency heatmap.

**Figure 2.**
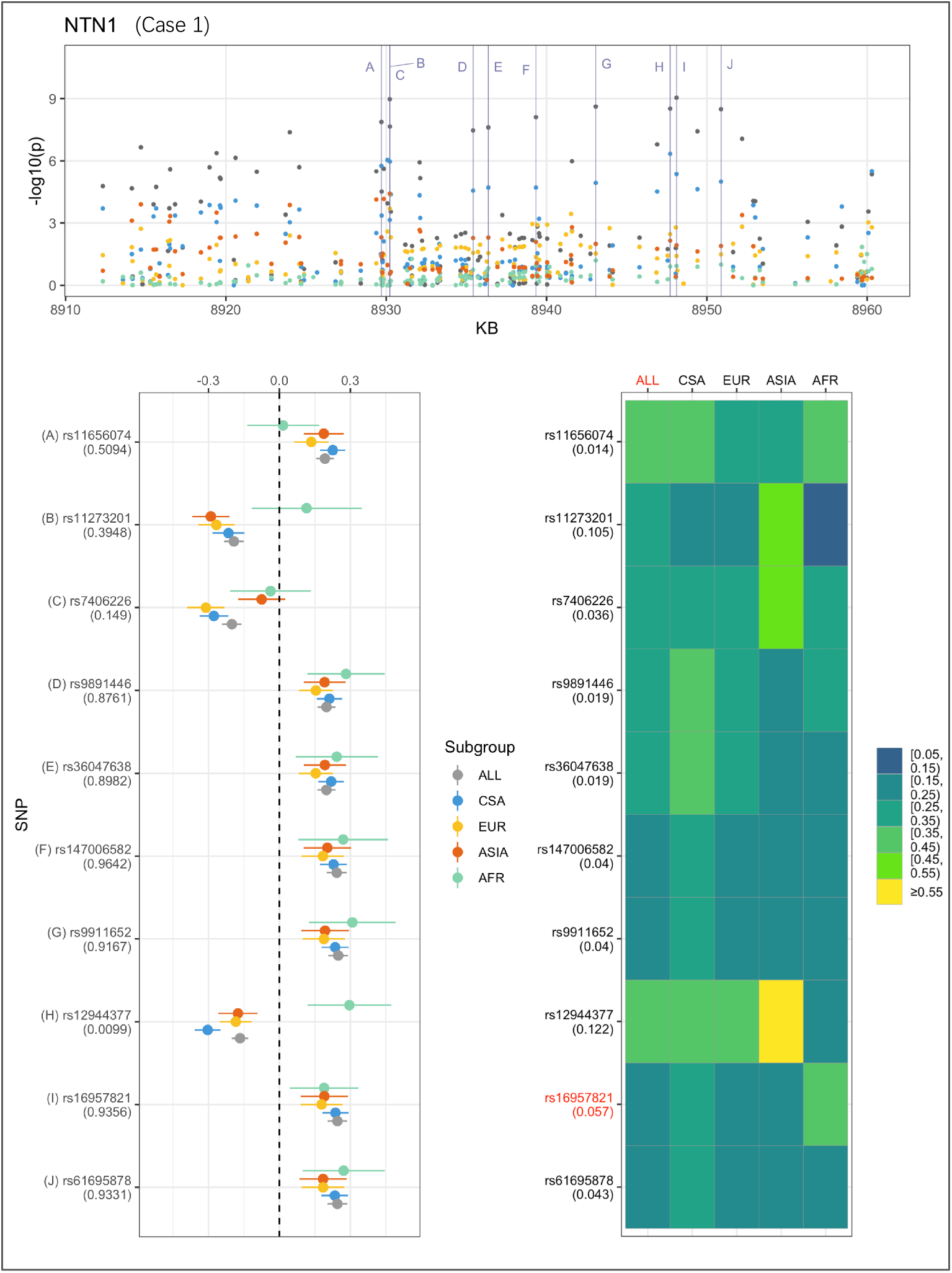
Association peak in 17p13.1 (*NTN1*) consistent with category 1. SNPs are ordered by base-pair position in lower panels; Q-statistic p-value noted under each SNP in forest plot of effect size; Wright’s F_ST_ value noted under each SNP name in heatmap of effect allele frequencies; lead variant and group with most significant p-value shown in red in heatmap.

**Figure 3.**
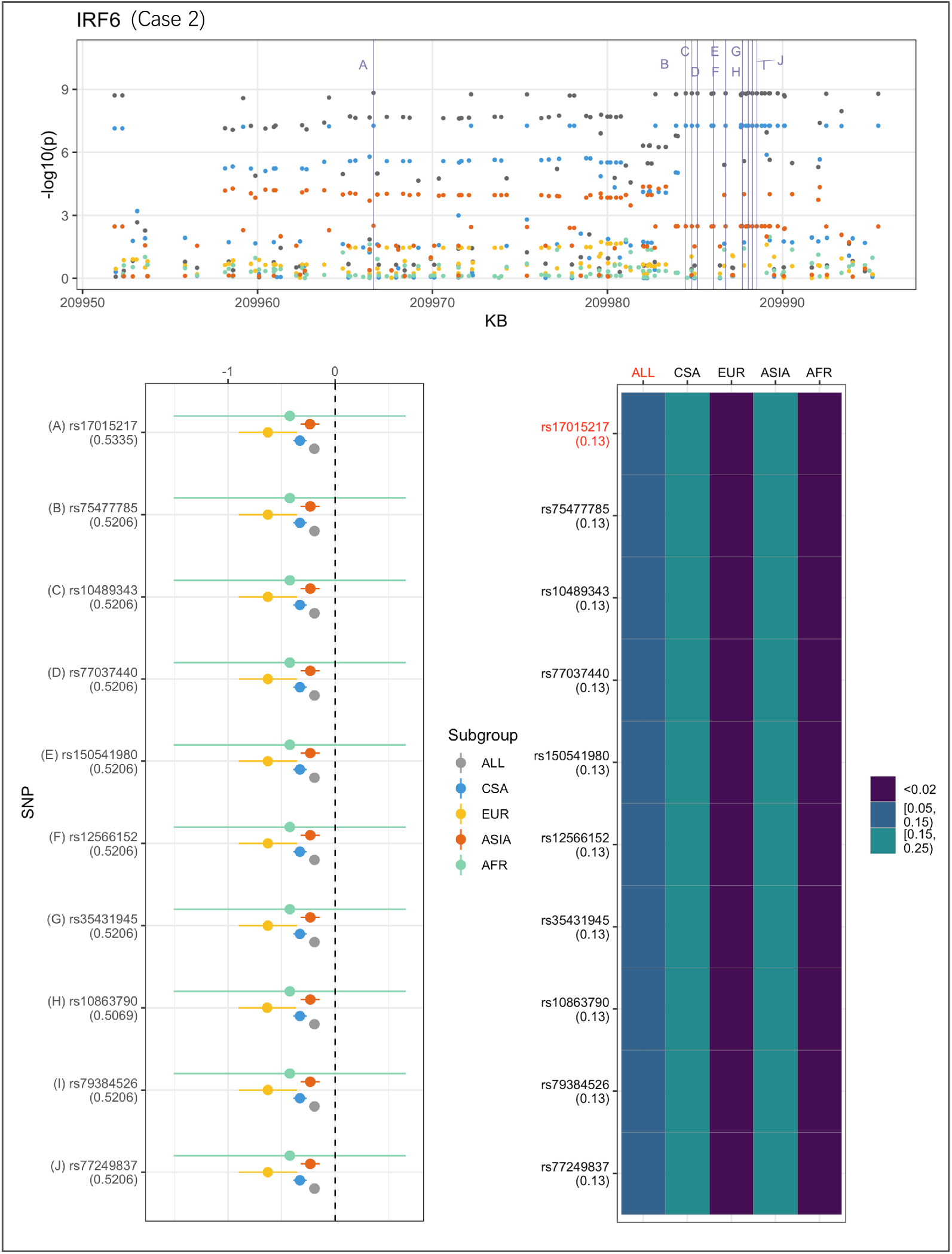
Association peak in 1q32.2 (*IRF6*) consistent with category 2. SNPs are ordered by base-pair position; Q-statistic p-value noted under each SNP in forest plot; lead variant and sample with highest p-value shown in red in heatmap and F_ST_ values noted under each SNP. The top 10 associated variants fall below our MAF filter in the EUR and ASIA groups.

**Figure 4.**
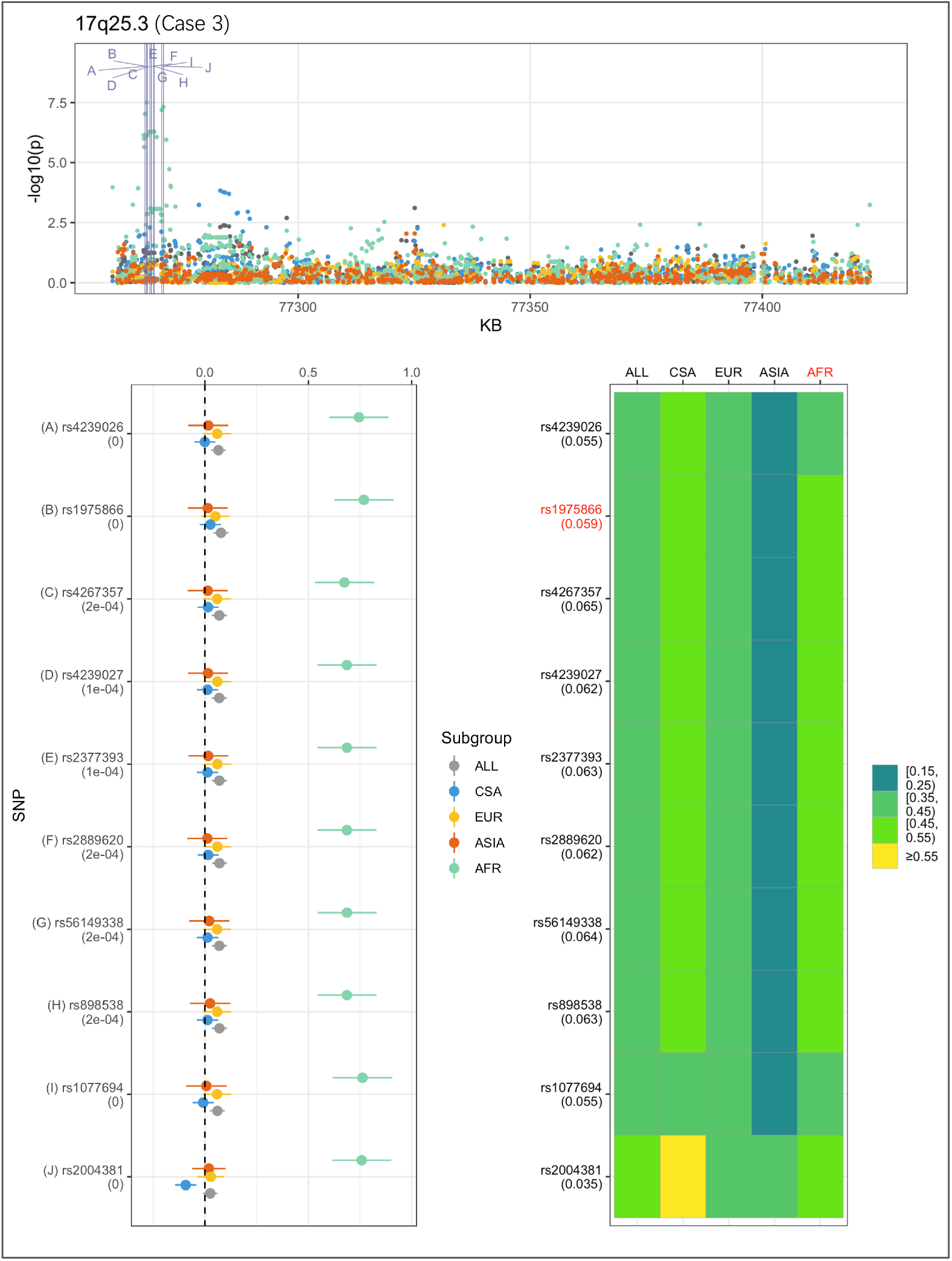
Association peak in 17q25.3 in the AFR group consistent with category 3. SNPs are ordered by base-pair position; Q-statistic p-value noted under each SNP in forest plot; lead variant and sample with highest p-value shown in red in heatmap, and F_ST_ values noted under each SNP.

In Figure 2, the largest sample, CSA produces the highest association p-values, followed by EUR and ASIA, and AFR. The effect sizes are very similar across the groups. The combined ALL sample shows the strongest association, and all four of the ancestry-based groups appear to contribute evidence for association at the 17p13.1 (*NTN1*) locus. This association conforms to the characteristics that describe category 1.

In figure 3, CSA and ASIA show elevated association p-values at the 1q32.2 (*IRF6*) locus, and are the major contributors to the significant association observed for the combined ALL sample, whereas EUR and AFR are weakly associated. The variants’ effect alleles are common in CSA and ASIA, and rare in the other two samples. The effect sizes are all similar, therefore, it is likely that the heterogeneity observed at this locus is due to difference in allele frequencies. The *IRF6* association conforms to category 2.

In Figure 4, at the 17q25.3 locus, the effect alleles are common in all four groups, however, a significant association is observed only in AFR. This is consistent with the small effect sizes observed in all groups except AFR. The 17q25.3 locus conforms is consistent with category 3.

## 4 Discussion

The current study extends previously published GWAS studies by using a substantially larger study sample than the previous GWAS study that included unrelated case trios, and singleton cases and controls. The larger sample size allowed us to identify three new potential loci that confer risk of CL/P, however, the addition of participants also contributed to added genetic heterogeneity, that diminished the strength of association in some cases.

One of the striking findings from OFC GWAs to date is the difference in association signals between populations. Although some loci such as 1q32 (IRF6) have been replicated in almost every population, most loci have not replicated between different populations. The lack of replication of associated loci between different populations can be attributed to differences in phenotype, sample size, or differences in allele frequency of the variants themselves. For example, the 8q24 locus is strongly associated in European populations but has not been detected in Asian populations because the risk alleles in Europeans have low frequencies in Asian populations, limiting the power to detect associations (Murray et al., 2012).

In our current study, we explored the genetic heterogeneity of cleft lip with or without cleft palate as a result of difference in ancestry, by running GWASs of CL/P within participants classified into four ancestry groups. As a result, we found that association outcomes differed substantially by ancestry. Our hypothesis was that variants controlling the risk of CL/P differs by ancestry, although these variants are distributed similarly. To test this hypothesis, we compared the main SNP effects of top associations in each locus to their observed allele frequencies. Our comparison showed evidence that some loci confer risk in every population, and the likelihood of observing an association is low in populations where these variants occur at a low frequency.

More importantly, we identified loci at which, the variants produced significantly different association outcomes, even though they were similarly distributed across the ancestry-based groups. It is likely that these loci do not modulate genetic risk of CL/P directly, but through gene-by-gene and gene-by-environment interactions.

An analysis of genetic diversity across the sites within each ancestry group showed that the ASIA and AFR groups are most diverse (median F_ST_ 0.02), indicating more distinct population admixture within those samples, while the EUR and CSA groups show lower diversity (median F_ST_ 0.003 and 0.009 respectively). It is likely that the genetic diversity of the ASIA sample was responsible for the lack of genome-wide significant or even suggestive associations, although the sample size is adequate (445 affected and 1,246 unaffected participants). The ASIA sample is geographically diverse, consisting of participants from China, India, and Philippines. The AFR and CSA groups are also geographically diverse (although CSA has a low F_ST_ genetic diversity coefficient), which could have led to weaker associations; there are only two genome-wide significant associations in CSA, even though this is our largest group consisting of 1,050 affected and 2,988 unaffected participants. In a previous whole genome sequence study that included only the CSA participants from Colombia, we observed a genome-wide significant association in chromosome 21q22.3 (Mukhopadhyay et al., 2020), but no corresponding association is seen either in this current study or the two previous studies that used CSA participants (Leslie et al., 2016; Leslie et al., 2017). The variants in the chromosome 21q22.3 in Colombian CL/P trios were observed to have similar allele frequencies across the subpopulations that make up the CSA group, but yielded no significant association when non-Colombian CSA participants were analyzed separately. That locus now appears to fit the characteristics of our current study’s category 3 loci.

Another contributing factor to the genetic heterogeneity is due to our phenotype itself. CL/P combines the two cleft subtypes: cleft lip only, and cleft lip with cleft palate. It has been shown by previous studies that the two subtypes are etiologically different (Carlson et al., 2017; Carlson et al., 2019). In the POFC multiethnic study sample, the frequency of these two subtypes differ among the sub-populations present, which can further reduce the power to detect association.

The exploration of the genetic differences between cleft subtypes is the focus of our ongoing investigation. This current study on the characterization of ancestry-level differences is an important step towards a more fine-grained and detailed investigation into the genetic heterogeneity of OFCs.

## Data Availability

Genetic and clinical data used in this study is publicly available from dbGaP and the FaceBase repository via the URLs indicated below.

http://www.ncbi.nlm.nih.gov/projects/gap/cgi-bin/study.cgi?study_id=phs000774.v2.p1

https://www.facebase.org/chaise/record/#1/isa:dataset/RID=1-50DT?pcid=recordset&ppid=1nel29se1r792mbi23uu1r6t

## 6 Author Contributions

NM, EF, EJL, and MLM were responsible for the study design and manuscript preparation. NM performed the data analysis. LM-U, GW, LV-R, CRM, CRP, FD, KC, FAP, IMO, JTH, CJB, AB, WLA and ARV contributed participant data to this study. All the authors reviewed the manuscript.

## 8 Acknowledgements

Many thanks to the participant families worldwide, without whom this research would not have been possible. Special thanks to Dr. Eduardo Castilla and Dr. Andrew Czeizel, and to the devoted staff at the many recruitment sites.

